# Genome-wide Associations, Polygenic Risk, and Mendelian Randomization Reveal Limited Interactions between John Henryism and Cynicism

**DOI:** 10.1101/2022.12.12.22283345

**Authors:** Richard R. Chapleau

## Abstract

Chronic occupational stress and an individual’s reaction to that stress often lead to burnout syndrome. We sought to use genetics to evaluate what, if any, interactions exist between John Henryism (JH) and cynicism in hopes of clarifying holistic risk factors of burnout syndrome. We performed genome-wide association studies in a discover phase with 1,852 samples and validated associations in a replication phase of 465 samples, both from the CARDIA study, and used supervised machine learning to developing genetic risk algorithms. We identified 933 genetic associations and developed a classification algorithm for high cynicism using machine learning with areas under the receiver operator characteristics curve greater than 0.7. We found significant genetic components to these traits but no evidence of an interaction between JH and cynicism, so while there may be a genetic risk component, JH does not appear to contribute to burnout risk.

## Introduction

First described formally by Freudenberger (1974) and later expanded upon by Maslach and Jackson (1981), burnout syndrome is generally considered to be a response to long-term occupational stress (Güler et al., 2019). The most widely used definition of burnout syndrome is a 3-component syndrome comprised of emotional exhaustion, depersonalization or cynicism, and feelings of low professional efficacy or personal accomplishment (Jackson et al., 1986). High levels of burnout syndrome components have been associated with increased disease prevalence including heightened risk of cardiovascular disease (Honkonen et al, 2006; Melamed et al., 1992), impaired cognitive function (Koutsimani et al., 2021), increased sleep disorders (Amaral et al., 2021; Stewart & Arora, 2019), and even type II diabetes and hyperlipidemia (Melamed et al., 2006; Shirom et al., 1997). Clearly, the impacts of burnout syndrome extend beyond just performance in the workplace and can adversely affect health, well-being, and quality of life.

Another response to chronic social stressors is John Henryism, a coping mechanism by which an individual exerts increased effort to overcome stress (Rolle et al., 2021). As with burnout syndrome, the effects of John Henryism affect both the individual’s health and the workplace. Also like the effects of burnout syndrome, John Henryism can have a negative impact on cardiovascular health and increased hypertension (James, 1983; James et al., 1994; Lebrón et al., 2015) and on increased rates of alcoholism (Vargas et al., 2020). Paradoxically, high levels of John Henryism have been associated with lower levels of depressive symptoms in African Americans of varying socioeconomic status, suggesting a protective effect on mental health (Robinson & Thomas Tobin, 2021). These findings suggest a nuanced interaction between John Henryism and overall health and well-being.

The literature on the genetic contribution to burnout syndrome and John Henryism is limited. Preliminary work regarding the heritability of cynicism (one of the three components of burnout syndrome) was reported in the late 1980’s to early 1990’s and revealed mixed results. One twin study of cynical hostility revealed a non-significant genetic component (Cates et al., 1993). In contrast, three other twin studies using the Cook Medley Hostility Scale and the MMPI showed a significant heritability (Carmelli et al., 1988; Rose, 1988; Smith et al., 1991). The genetic heritability of John Henryism also has limited reports, with one estimate suggesting up to 35% of the variability is explained by genetic factors (Wang et al., 2005; Whitfield et al., 2006). To the best of our knowledge, there have not been any genome-wide or candidate gene studies performed to assess the genetic contribution to these traits.

Here we report the results of our study investigating the relationship between John Henryism and cynicism. We hypothesized that John Henryism would exert a causal influence on cynicism. We came to this hypothesis because of the nature of John Henryism as a coping mechanism for dealing with discriminatory acts and the skepticism and negative view of others inherent to cynicism. We tested this hypothesis through the statistical approach called Mendelian randomization (Hemani et al., 2018), where genetic associations with John Henryism are considered for their independent influence on cynicism. In order to take this approach, we first performed a genome-wide association study (GWAS) to identify genetic variables for John Henryism and cynicism independently. We also extended those GWAS results to develop polygenic risk scores (risk scores considering multiple genetic variants) for each trait and assessed if higher genetic risk in one trait correlated to higher levels of the other trait. To our knowledge, this study is the first to report genetic associations with any of the three outcomes.

## Methods

This study was reviewed and approved by WCG IRB (Study number 1332892) for human subjects research oversight. All data were obtained from the National Institutes of Health’s Database of Genotypes and Phenotypes (dbGAP). We used data from the Coronary Artery Risk Development in Young Adults (CARDIA) Study (dbGAP study accession phs000285) (Friedman et al., 1988). The CARDIA cohort study design was a longitudinal study, but we used the data in a retrospective fashion. We followed the STREGA guidelines available from the EQUATOR network (Little et al., 2009). The guideline table with annotations regarding how we addressed each point is available as supplementary material.

### Psychological trait definitions

John Henryism (JH) was measured by the 12-item John Henryism Active Coping Scale (JHAC), and responses were reverse-coded. JH was calculated as a mean of the responses. As our goal was to observe the effect of having above-average JH, we defined a dichotomous variable of JH as individuals with scores above the median (49) as having high JH (James et al., 1983; Lebrón et al., 2015). Individuals with scores at or below the median were considered average or below average JH. We also performed tests using the JH score as a continuous, mean-centered variable. The JH data were obtained from dbGAP accession number phv001133534.v2.p2.c1. Cynicism and cynical distrust (CD) were measured as 12- and 8-item subscales of the Cook-Medley Hostility Scale (CMHS), respectively (Barefoot et al., 1991; Greenglass & Julkunen, 1991; Wong et al., 2013). Mean-centered continuous variables and median-adjusted dichotomous variables were created for cynicism and CD in the same manner as for JH, resulting in four CMHS-derived variables (continuous cynicism, high/low cynicism, continuous CD, and high/low CD). The CMHS data were obtained from dbGAP accession number phv00113478.v2.p2.c1. As not all participants completed all questionnaires, missing data were filled in as the mean. Processed data were then split into a validation set (20%, *n* = 623) and the remainder were used for training (Figure 1). Statistical analyses were performed in R version 4.2.0.

**Figure 1.**
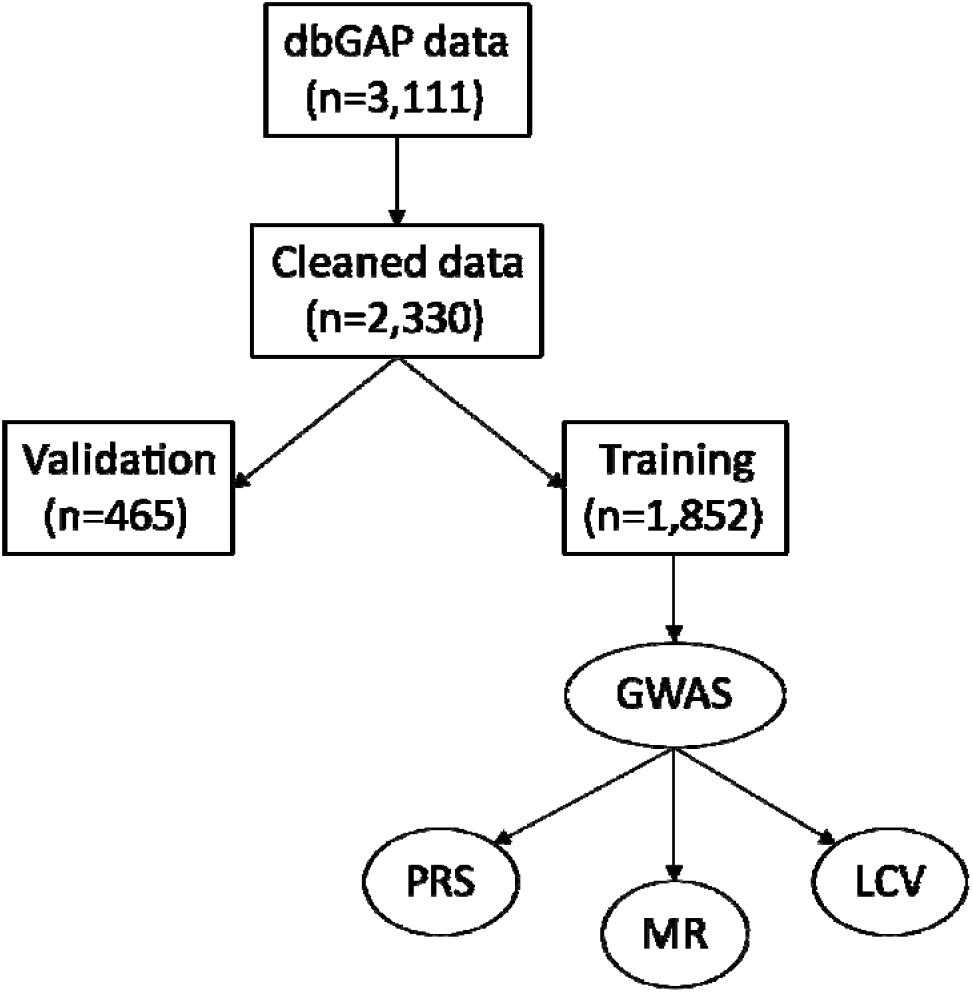
Flow diagram for data processing. Rectangles are datasets with sample size in parentheses and ovals are processes. dbGAP = Database of Genotypes and Phenotypes; GWAS = genome-wide association studies; LCV = latent causal variable; MR = Mendelian randomization; PRS = polygenic risk score.

### Power analysis

We performed a power analysis for our study to determine if the sample sizes available in the CARDIA cohort were sufficient to identify significant genetic associations. We assumed a 10% effect allele frequency (EAF) in the control population, 20% type-II error rate (beta) and 5% type-I error rate (alpha). We used the observed case-control ratios for each condition to calculate the statistical power. We report the results of the analysis as the minimum EAF in the case population to achieve 80% power in Table 1 alongside the cohort characteristics.

**Table 1.** Breakdown of samples by dichotomous traits.

|  | John Henryism | Cynicism | Cynical Distrust |
| --- | --- | --- | --- |
| Training High (%) | 877 (47.4%) | 724 (39.1%) | 680 (36.7%) |
| Training Not High | 975 | 1,128 | 1,172 |
| Case:Control Ratio | 1:1.1 | 1:1.6 | 1:1.7 |
| Minimum EAF | 14.5% | 15.7% | 16.0% |
| Validation High (%) | 246 (52.9%) | 180 (38.7%) | 171 (36.8%) |
| Validation Not High | 219 | 285 | 294 |

*EAF = effect allele frequency*

### Genetic data pre-processing

Microarray (Affymetrix Genome-Wide Human SNP 6.0 Array) genotype data were obtained from the CARDIA study (dbGAP accession numbers phg000092.v2 and phg000098.v2). Genetic data were pre-processed to ensure uniformity from the original plink data. Briefly, the binary plink filesets were merged and filtered for autosomal genotypes with less than 10% missing genotype calls (sample or locus), a minor allele frequencies threshold was set at 1%, and a Hardy-Weinberg equilibrium threshold of .0001 was used for filtering out spurious variants. After pre-processing, they were split into the training and validation sets using the sample identifiers defined in the phenotype data splitting.

### Genome-wide association studies

We conducted GWAS using plink 1.9 evaluating only the total scores (not scores for the questionnaire responses) and high/low status for each trait, totaling 6 phenotypes. We used the following command for the association:

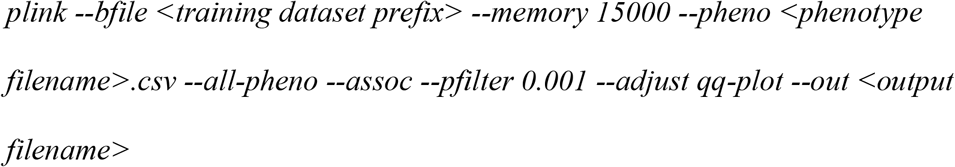

For assessing the replication of associations in continuous variables traits, we repeated the GWAS in plink restricting the input variants to only those candidates identified in the discovery phase. For dichotomous traits, the set of candidate SNPs identified as significantly associated with the trait (*p* < 5x10^−8^) or suggestively associated (*p* < 5x10^−6^) were then evaluated by chi-square test using the test dataset. Additionally, each analysis was repeated conditioning upon the highest associated SNP.

### Polygenic risk score calculation

Our method of PRS development was using the machine learning package scikit-learn (Pedregosa et al., 2011). We used four supervised classifiers (Ridge, multi-layer perceptron (MLP), random forest (RFC), and k-nearest neighbors (KNN)) and iterated through the relevant parameter space for each classifier (e.g., number of neighbors for KNN). The classifiers were trained on the training set (*n* = 1852) and validated using the test dataset (*n* = 465). Each classifier was evaluated using the area under the receiver operator characteristics (ROC) curve (AUC) and the model with the best AUC was saved for each classifier. Finally, a ROC curve comparing the best models of each classifier was created.

### Mendelian randomization

We performed two-sample Mendelian randomization (2SMR) estimates using the TwoSampleMR R package (Hemani et al., 2018). We used the MR Egger regression (Bowden et al., 2015), inverse variance weighted (IVW) estimator (Burgess et al., 2013), weighted median estimator (Bowden et al., 2016), and Wald ratio estimator (Teumer, 2018) algorithms. Instrumental variables (IV, SNPs associated with the exposure) were extracted by *p*-value thresholds 5x10^−6^ and 5x10^−8^. We excluded SNPs in strong linkage disequilibrium (LD) to reduce bias and used a clumping process with European samples from the 1,000 Genomes Project (*r*^2^ < .001, window size = 10,000). If SNPs identified in the exposure dataset were not in the outcome dataset, proxy SNPs in LD (*r*^2^ > .9) were used as instrumental variables. For the sensitivity analysis, we performed heterogeneity testing using Cochran’s Q and I^2^ analyses (Greco et al., 2015) and tested pleiotropy on the weighted median estimation results (Bowden et al., 2016).

## Results

### Characteristics of the dataset

The initial phenotype dataset consisted of 3,111 samples (Figure 1). The median scores for John Henryism, cynicism, and CD were 49, 6, and 3, respectively. Score ranges were 26 to 60 for JH, 0 to 13 for cynicism, and 0 to 8 for CD. There were 1497 (48.1%) samples with high JH scores (JH > 49), 1228 (39.5%) with high cynicism (> 6), and 1163 (37.4%) with high CD scores (>3). We found minimal correlation between John Henryism and cynicism or CD (Pearson *r* = .159 and .118, respectively). After splitting the data into training (80%) and validation (20%) sets, the median scores were 49, 6, and 3, respectively, for the training set and 50, 5, and 3, respectively, for the validation set. The percent of samples above the population median scores was 47.1%, 40.0%, and 37.8%, respectively, for the training set and 52.3%, 37.4%, and 35.6%, respectively, for the validation set. T-tests showed that the distribution of samples in the subsets was not statistically different from the original distribution (JH training vs. original *p* = .52; JH validation vs. original *p* = .12; cynicism training vs. original *p* = .48; cynicism validation vs. original *p* = .08; CD training vs. original *p* = .62; and CD validation vs. original *p* = .22). Evaluating the Shapiro-Wilk normality test in R reveals that none of the six variables are normally distributed (all have *p* < 2.2x10^−16^). The Shapiro-Wilk *W* constants for the distributions of quantitative scores were .980, .935, and .970 for John Henryism, CD, and cynicism, respectively, while the *W* constants for the dichotomous traits were .635, .615, and .622, respectively. The skewness of the three continuous traits was -0.511, 0.118, and 0.373, respectively, while the kurtosis values were 3.11, 1.01, and 2.18, respectively. From these results we see that the data are generally positively skewed and primarily platykurtic.

The merged genetic dataset contained 2,466 samples (1,162 from accession phg000092 and 1,441 from accession phg000098, with 137 overlapping between the two studies), of which 978 (42.1%) were male. The two datasets contained 909,662 and 720,622 markers, respectively. Following SNP filtering, there were 561,045 variants remaining (153,333 for missingness, 13,410 below 1% MAF, and 144,454 not passing Hardy-Weinberg filter). 136 samples were removed for having greater than 10% missing genotype calls, creating a final dataset of 2,330 samples. The total genotyping rate for the dataset was 99.4%. After splitting samples using the training and validation sets defined in the phenotype stage, there were 1,852 samples in the training set, 465 samples in the validation set, and 13 (0.6%) of the samples with genetic data did not have phenotype data (Table 1).

### GWAS Results

Our discovery phase GWAS analysis of 1,852 samples revealed 25 candidate variants with Bonferroni-corrected *p*-values below 5x10^−8^ associated with John Henryism, 28,926 candidates associated with cynicism, and 14,134 candidate associations with CD (Figure 2). For each of the candidate variants, we performed a second GWAS on the test sample set (*n* = 465) using only the candidate SNPs identified in the first analysis. We found that 2 SNPs replicated as associated with the quantitative John Henryism trait (*p* = .002), 727 replicated for association with cynicism (*p* = 1.73x10^−6^), and 204 replicated for association with CD (*p* = 3.54x10^−6^).

**Figure 2.**
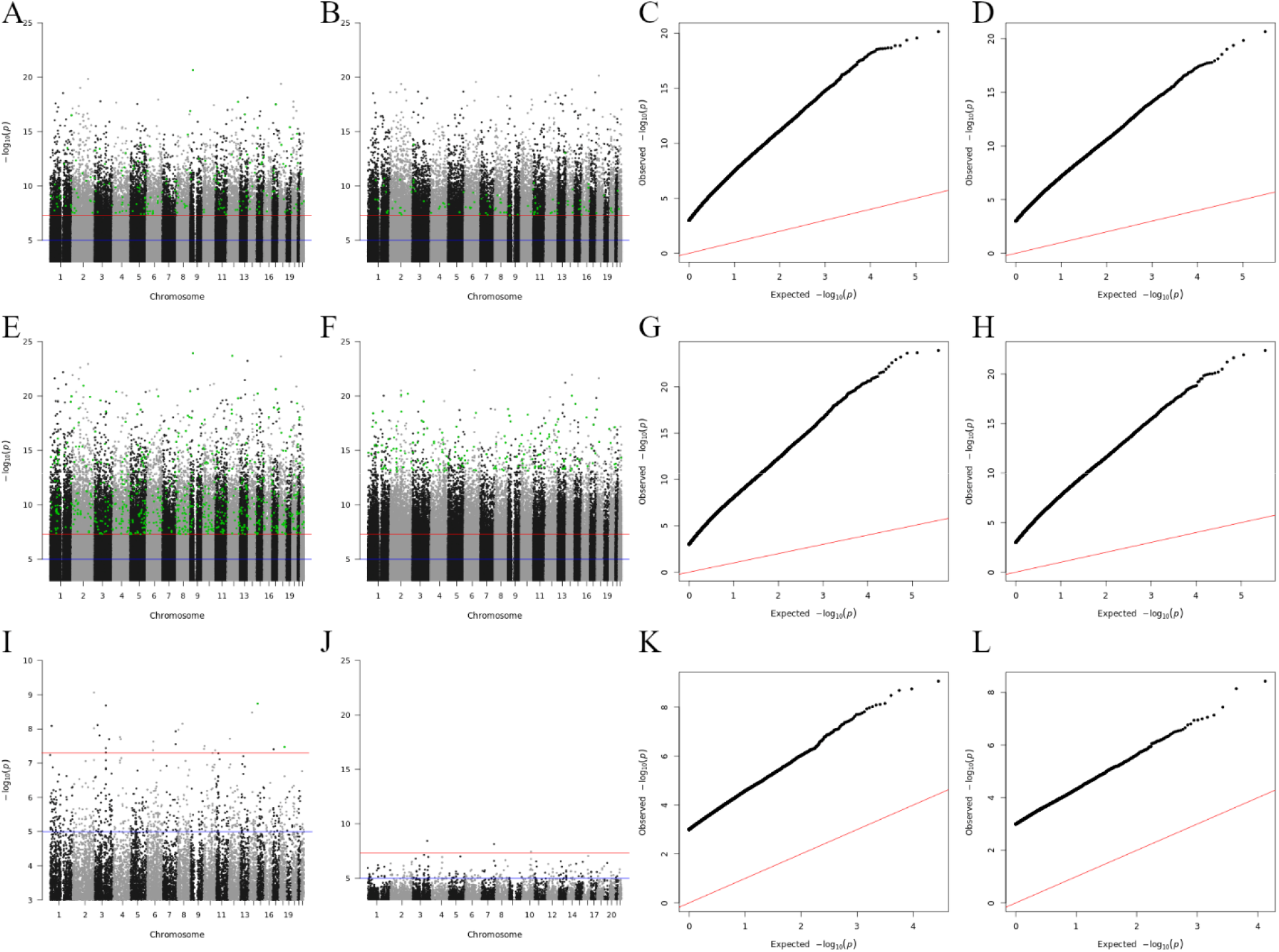
Manhattan (left six) and quantile-quantile (right six) plots of genome-wide associations for cynical distrust (A, B), cynicism (C, D) and John Henryism (E, F). Plots are shown for the continuous trait (A, C, and E) and the dichotomous trait (B, D, and F). Black dots = candidate associations from the training set (n=1852); green dots = associations replicated in the verification set (n=465); blue line = genome-wide suggestive association (P = 5x10-6); red line = genome-wide significant association (P = 5x10-8).

Similar to how we analyzed the continuous variables, we identified in the dichotomous variable analysis 3 candidate variants associated with high John Henryism, 708 associations with high cynicism, and 17,507 associations with high CD. After evaluating for replication of significance (*p* < .05) in the validation set of 465 samples, we found 0, 173, and 109 significantly associated variants that replicated in our test set for John Henryism, cynicism, and CD, respectively. Of the 173 replicated variants associated with cynicism, 19 were located across 9 distinct loci (defined as within a 250,000-base pair window on the same chromosome) and the other 154 were distinct variants. We also observed that 79 of the 173 high cynicism associated variants were also associated with the quantitative trait, while none of the high CD associated variants were present in the quantitative trait CD list. Lists of all candidate and replicated variants are available as from the corresponding author upon reasonable request.

### ML-based polygenic risk scores

We used the replicated SNPs for each condition cynicism and CD to evaluate the predictive capability for each, addressing the question of whether genetic variants associated with high John Henryism, for example, were predictive of high cynicism. As no SNPs replicated for high John Henryism, we used the candidate SNPs. Using the scikit-learn software package, we performed the nine tests for each input/output combination (e.g., cynicism vs. CD, cynicism, and John Henryism) with the four classification methods. Our results (Figure 3, Table 2) reveal that PRS algorithms based on genetic variants associated high cynicism are predictive of high cynicism (AUC range = .696-.732) and high CD (AUC range = .652-.684), whereas algorithms trained on genetic markers of high JH or high CD are not predictive for any trait. These classifiers would be considered to be acceptable predictors (Mandrekar, 2010) with AUC values near 0.7, these results show that cynicism and CD are genetically related, reinforcing the psychological relationship, and that JH is a distinct trait deriving from different genetic contributions.

**Figure 3.**
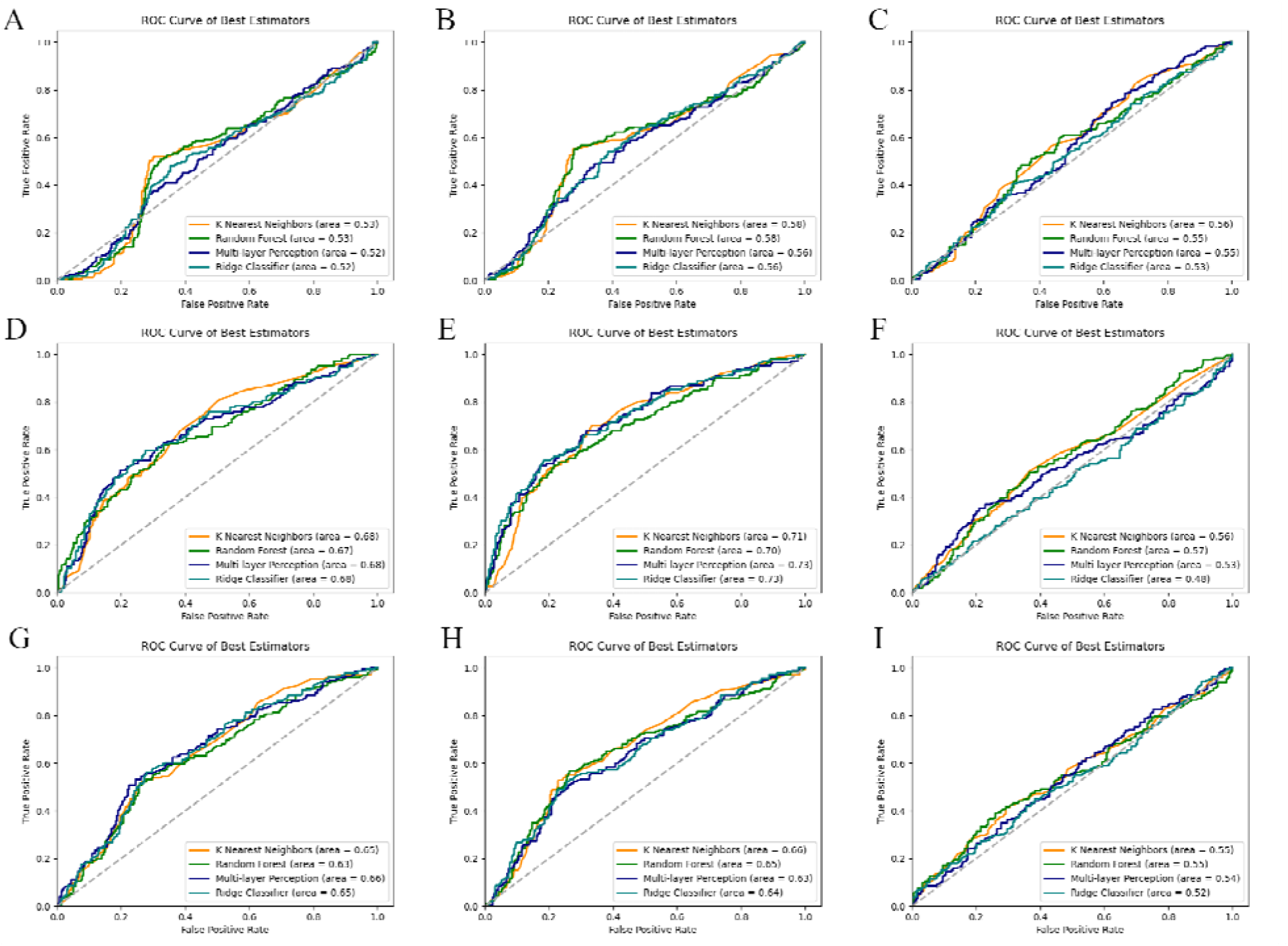
Receiver operator characteristics (ROC) curves for genetic classification algorithms for high cynical distrust (A-C), high cynicism (D-F), and high John Henryism (G-I). Each panel shows four predictors (orange = K-nearest neighbors, green = random forest, navy = multi-layer perceptron, teal = ridge classification) measured for its ability to predict cynical distrust (A, D, G), cynicism (B, E, H) or John Henryism (C, F, I).

**Table 2.** AUC values for ML-based polygenic risk algorithms.

| Input vs. Output Pair | Ridge | MLP | RFC | KNN |
| --- | --- | --- | --- | --- |
| JH vs. JH | 0.524 | 0.542 | 0.545 | **0.550** |
| JH vs. cynicism | 0.638 | 0.634 | 0.649 | **0.660** |
| JH vs. CD | 0.653 | **0.656** | 0.651 | 0.630 |
| Cynicism vs. JH | 0.482 | 0.533 | **0.565** | 0.563 |
| Cynicism vs. cynicism | **0.732** | 0.728 | 0.696 | 0.712 |
| Cynicism vs. CD | 0.681 | 0.676 | 0.671 | **0.684** |
| CD vs. JH | 0.526 | 0.545 | 0.546 | **0.563** |
| CD vs. cynicism | 0.563 | 0.556 | 0.577 | **0.580** |
| CD vs. CD | 0.516 | 0.516 | 0.526 | **0.533** |

*JH = John Henryism, CD = cynical distrust, MLP = multi-layer perceptron, RFC = random forest classifier, KNN = k-nearest neighbors. Bold values are greatest area under the receiver operator characteristic curve for the input-output pairing*

### Two sample Mendelian randomization

Our two sample Mendelian randomization analyses using the GWAS summary statistics previously described revealed significant causal relationships between all three traits (Figure 4). The odds ratios (Figure 4A) and beta coefficients (Figure 4B) for the relationships were highly similar, with odds ratios ranging from 2.0 (for the causal effect of John Henryism on cynicism and the causal effect of cynical distrust on John Henryism) to 3.3 (for the inverse effects), with the cynical distrust/cynicism relationship near 2.6. The MR comparisons involved 4,680 variants for the John Henryism/cynicism relationship, 4,262 variants for the John Henryism/cynical distrust relationship, and 131,185 variants for the cynical distrust/cynicism relationship. We performed pleiotropy and heterogeneity tests to assess the sensitivity of the MR estimates. We found significant pleiotropy in all cases, with the Egger intercepts significantly different from zero (Figure 4C) (Burgess & Thompson, 2017).

**Figure 4.**
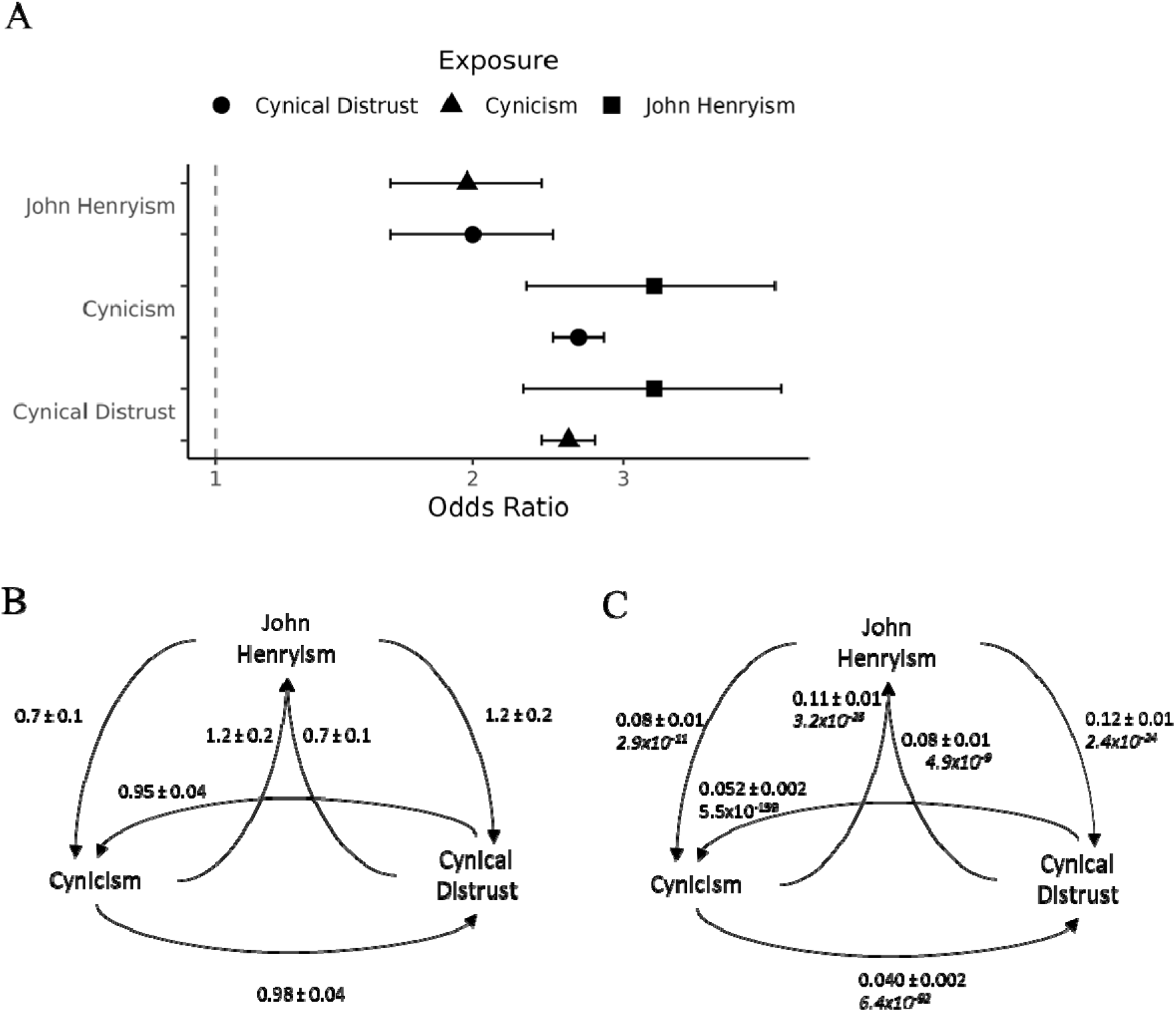
Results from Mendelian randomization. A) Average odds ratios and 95% confidence intervals of the causal relationships between traits as determined by Mendelian randomization. B) Average beta coefficients and standard errors from four MR methods. C) Egger intercepts, standard errors, and P-values (italic) from pleiotropy analyses.

## Discussion

Our results from genetic analyses suggest that there is not a significant relationship between John Henryism and cynicism. These observations are consistent with previously published literature using only psychometric approaches. Adams and co-workers (1999) demonstrated that John Henryism scores were not significantly correlated with the Cook Medley Hostility Scale (Ho), of which we used the cynicism subscale herein. In contrast, a large study investigating the effects of various psychosocial factors on chronic kidney disease found that John Henryism and hostility exhibited inverse risk factor loading (Lunyera et al., 2018). Finally, initial reports from the CARDIA study suggest a weak but significant correlation between hostility and effortful coping (*r* = 0.14, *p* < 0.05), especially among younger, less educated individuals that were more likely to consume alcohol and be smokers (Albanese et al., 2016).

Although our findings do not substantiate a relationship between John Henryism and cynicism, and subsequently burnout syndrome, they do suggest that there is a genetic component to both outcomes. Research among the general population has shown that hostility is an independent risk factor for cardiovascular disease (Matthews et al., 2004), which has also been reinforced with evidence in population-specific work in African Americans (Bhavsar et al., 2021). Our results suggest that a significant predisposition to hostility and cynicism, when coupled with chronic occupational stress and hostility, could lead to elevated levels of burnout and significant long-term health decline.

Our assessment of causal effects revealed significant pleiotropy in the relationships, suggesting that any interaction which may exist is indirect. Our results do not support a causal effect, but neither do they provide sufficient evidence to refute any such relationship. Indeed, our assessment using multiple MR estimators provides limited support that there is a relationship between these traits, but that it such an interaction may not be mediated by genetics. We used the MR Egger estimate to evaluate violations of the instrumental variable assumptions. We found that these assumptions are not valid in our analysis (the Egger intercepts were significantly non-zero). However, the magnitude of the impact of any deviations must be small as none of the six beta coefficients determined by the MR Egger method were outliers from the other four estimates (i.e., all MR Egger estimates were within the 95% confidence interval of the mean observed beta coefficient). Therefore, it appears that using the MR Egger estimator did not introduce additional bias or increase the Type I error rate (Burgess & Thompson, 2017).

The pleiotropic analysis suggests that there are confounders involved in the interaction and, therefore, another limitation of our study was that we did not consider covariates in the initial analysis. The CARDIA study included many demographic and phenotypic datasets from which covariates could be identified. As the outcomes we measured tend to change with age (Albanese et al., 2016), at least age at the time the questionnaire could be used as a future covariate. Indeed, the weak correlations reported by Albanese were largely derived from stratified data. While it was beyond the scope of our initial view of the interactions between genetics, John Henryism, and cynicism to assess covariates, additional covariates could include gender, race, or personality, among others.

One major limitation in this study is that the sample size is relatively small for a large-scale genomics study. Our discovery phase was comprised of only 1,852 samples and our replication phase consisted of another 465 samples. While an *a priori* power analysis revealed that these sample sizes should have been powered to identify significant associations, it would be prudent to repeat these observations in larger populations to confirm which, if any, variants remain significantly associated with John Henryism, cynicism, and/or cynical distrust. Due to this sample size limitation, the generalizability of our results should be considered alongside larger scale studies, especially with regards to burnout syndrome, for which not much is published in the genetic literature.

Analyzing the qq-plots reveals that there is a significant deviation from normality in all of the datasets (Figure 2). Considering the non-normal distribution of the data and the large sample distribution within the dataset, this deviation from normality was expected (both Cook-Medley and John Henryism scores were previously acknowledged to have skewed distributions) (Albanese, 2016). What was unexpected was the relatively large number of significantly associated variants for cynicism and cynical distrust found in both the discovery and replication phases. Viewed alone, the large number of associated variants is cause for skepticism in the results. However, when taken into consideration along with the predictive ability of ML algorithms (Figure 3) developed based upon these replicated variants, the evidence for a significant genetic contribution to cynicism and cynical distrust becomes stronger.

In conclusion, our results suggest that high levels of cynicism and cynical distrust have a significant genetic component and there may some genetic component to levels of John Henryism. This genetic component of cynicism and cynical distrust appears to have some common effects as polygenic risk scores developed to classify individuals with high scores in one trait are reasonably effective at classifying individuals with the other trait (AUC > .6). Our results are insufficient to determine if this correlation is based upon causation, however, as there is a significant amount of observed pleiotropy in the MR analysis, suggesting the existence of confounding variables.

## Data Availability

Genotype and phenotype data that support this study can be obtained from dbGAP using accession numbers phs000285.v3.p2, pht001579.v2.p2, pht001634v2.p2, pht001786.v2.p2, phg000092.v2, and phg000098.v2. GWAS summary statistics based created from GWAS on the training dataset are unavailable for public posting, per the CARDIA data use agreement. The summary statistics, ML code and models are available from the authors upon reasonable request.

## Acknowledgments

The authors wish to thank Mr. Billy Thompson, Ms. CharLee Martin, Mr. Eric Rigby, and Mr. Warren Fridy for their assistance in coordinating data access and security. The authors also wish to thank the participants and the original depositors of the CARDIA study.

## Declarations

### Funding

No explicit funding was received for this study.

### Conflicts of Interest

Richard Chapleau is the Principal Geneticist of NeuroStat Analytical Solutions, a for-profit company.

### Ethics Approval

This study was reviewed and approved by WCG IRB (Study number 1332892) for human subjects research oversight.

### Consent to Participate

Not applicable Consent for Publication: Not applicable

### Code Availability

Not applicable

### Authors’ Contributions

All aspects of this work were performed by RRC.

## References

Adams JH, Aubert RE, Clark VR. The relationship among John Henryism, hostility, perceived stress, social support, and blood pressure in African-American college students. Ethn Dis. 1999 Autumn;9(3):359–68. PMID: 10600058.

Albanese E, Matthews KA, Zhang J, Jacobs DR Jr, Whitmer RA, Wadley VG, Yaffe K, Sidney S, Launer LJ. Hostile attitudes and effortful coping in young adulthood predict cognition 25 years later. Neurology. 2016 Mar 29;86(13):1227–34. doi:10.1212/WNL.0000000000002517. Epub 2016 Mar 2. PMID: 26935891; PMCID: PMC4818565.

Amaral KV, Galdino MJQ, Martins JT. Burnout, daytime sleepiness and sleep quality among technical-level Nursing students. Rev Lat Am Enfermagem. 2021 Oct 29;29:e3487. doi: 10.1590/1518-8345.5180.3487. PMID: 34730763; PMCID: PMC8570253.

Barefoot JC, Peterson BL, Dahlstrom WG, Siegler IC, Anderson NB, Williams RB Jr. Hostility patterns and health implications: correlates of Cook-Medley Hostility Scale scores in a national survey. Health Psychol. 1991;10(1):18–24. doi: 10.1037//0278-6133.10.1.18. PMID: 2026126.

Bhavsar NA, Davenport CA, Yang LZ, Peskoe S, Scialla JJ, Hall RK, Tyson CC, Strigo T, Sims M, Pendergast J, Curtis LH, Boulware LE, Diamantidis CJ. Psychosocial determinants of cardiovascular events among black Americans with chronic kidney disease or associated risk factors in the Jackson heart study. BMC Nephrol. 2021 Nov 11;22(1):375. doi: 10.1186/s12882-021-02594-6. PMID: 34763649; PMCID: PMC8582093.

Bowden J, Davey Smith G, Haycock PC, Burgess S. Consistent Estimation in Mendelian Randomization with Some Invalid Instruments Using a Weighted Median Estimator. Genet Epidemiol. 2016 May;40(4):304–14. doi: 10.1002/gepi.21965. Epub 2016 Apr 7. PMID: 27061298; PMCID: PMC4849733.

Bowden, J., Davey Smith, G. and Burgess, S. (2015). Mendelian randomization with invalid instruments: Effect estimation and bias detection through Egger regression. Int. J. Epidemiol. 44 512–52

Bowden, J., Davey Smith, G., Haycock, P. C. and Burgess, S. (2016). Consistent estimation in Mendelian randomization with some invalid instruments using a weighted median estimator. Genet. Epidemiol. 40 304–314.

Burgess S, Thompson SG. Interpreting findings from Mendelian randomization using the MR-Egger method. Eur J Epidemiol. 2017 May;32(5):377–389. doi: 10.1007/s10654-017-0255-x. Epub 2017 May 19. Erratum in: Eur J Epidemiol. 2017 Jun 29;: PMID: 28527048; PMCID: PMC5506233.

Burgess, S., Butterworth, A. and Thompson, S. G. (2013). Mendelian randomization analysis with multiple genetic variants using summarized data. Genet. Epidemiol. 37 658–665

Carmelli D, Rosenman RH, Swan GE. The Cook and Medley HO scale: a heritability analysis in adult male twins. Psychosom Med. 1988 Mar-Apr;50(2):165–74. doi: 10.1097/00006842-198803000-00006. PMID: 3375406.

Cates DS, Houston BK, Vavak CR, Crawford MH, Uttley M. Heritability of hostility-related emotions, attitudes, and behaviors. J Behav Med. 1993 Jun;16(3):237–56. doi: 10.1007/BF00844758. PMID: 8350340.

Freudenberger HJ. Staff burnout. J Soc Issues. 1974;30:159–65.

Friedman GD, Cutter GR, Donahue RP, et al. CARDIA: study design, recruitment, and some characteristics of the examined subjects. J Clin Epidemiol 1988;41:1105–1116.

Greco M FD, Minelli C, Sheehan NA, Thompson JR. Detecting pleiotropy in Mendelian randomisation studies with summary data and a continuous outcome. Stat Med. 2015 Sep 20;34(21):2926–40. doi: 10.1002/sim.6522. Epub 2015 May 7. PMID: 25950993.

Greenglass ER, Julkunen J. Cook-Medley hostility, anger, and the Type A behavior pattern in Finland. Psychol Rep. 1991 Jun;68(3 Pt 2):1059–66. doi: 10.2466/pr0.1991.68.3c.1059. PMID: 1924607.

Güler Y, Şengül S, Çaliş H, Karabulut Z. Burnout syndrome should not be underestimated. Rev Assoc Med Bras (1992). 2019 Nov;65(11):1356–1360. doi: 10.1590/1806-9282.65.11.1356. PMID: 31800896.

Hemani, G., Zheng, J., Elsworth, B., Wade, K., Haberland, V., Baird, D., Laurin, C., Burgess, S., Bowden, J., Langdon, R., Tan, V. Y., Yarmolinsky, J., Shibab, H., Timpson, N., Evans, D., Relton, C., Martin, R., Davey Smith, G., Gaunt, T., Haycock, P., & The MR-Base Collaboration. (2018). The MR-Base platform supports systematic causal inference across the human phenome. eLife, 7(), e34408. https://doi.org/10.7554/eLife.34408

Honkonen T, Ahola K, Pertovaara M, Isometsä E, Kalimo R, Nykyri E, Aromaa A, Lönnqvist J. The association between burnout and physical illness in the general population--results from the Finnish Health 2000 Study. J Psychosom Res. 2006 Jul;61(1):59–66. doi: 10.1016/j.jpsychores.2005.10.002. PMID: 16813846.

Jackson, S. E., Schwab, R. L., & Schuler, R. S. (1986). Toward an understanding of the burnout phenomenon. Journal of Applied Psychology, 71(4), 630–640. https://doi.org/10.1037/0021-9010.71.4.630

James SA, Hartnett SA, Kalsbeek WD. John Henryism and blood pressure differences among Black men. J Behavioral Med. 1983;6(3):259–278.

James SA. John Henryism and the health of African-Americans. Cult Med Psychiatry. 1994;18:163–82.

Koutsimani P, Montgomery A, Masoura E, Panagopoulou E. Burnout and Cognitive Performance. Int J Environ Res Public Health. 2021 Feb 22;18(4):2145. doi: 10.3390/ijerph18042145. PMID: 33671754; PMCID: PMC7926785.

LeBrón AM, Schulz AJ, Mentz G, White Perkins D. John Henryism, socioeconomic position, and blood pressure in a multi-ethnic urban community. Ethn Dis. 2015 Winter;25(1):24–30. PMID: 25812248; PMCID: PMC4385581.

Little J, Higgins JP, Ioannidis JP, Moher D, Gagnon F, von Elm E, Khoury MJ, Cohen B, Davey-Smith G, Grimshaw J, Scheet P, Gwinn M, Williamson RE, Zou GY, Hutchings K, Johnson CY, Tait V, Wiens M, Golding J, van Duijn C, McLaughlin J, Paterson A, Wells G, Fortier I, Freedman M, Zecevic M, King R, Infante-Rivard C, Stewart A, Birkett N; STrengthening the REporting of Genetic Association Studies. STrengthening the REporting of Genetic Association Studies (STREGA): an extension of the STROBE statement. PLoS Med. 2009 Feb 3;6(2):e22. doi: 10.1371/journal.pmed.1000022. PMID: 19192942; PMCID: PMC2634792.

Lunyera J, Davenport CA, Bhavsar NA, Sims M, Scialla J, Pendergast J, Hall R, Tyson CC, Russell JSC, Wang W, Correa A, Boulware LE, Diamantidis CJ. Nondepressive Psychosocial Factors and CKD Outcomes in Black Americans. Clin J Am Soc Nephrol. 2018 Feb 7;13(2):213–222. doi: 10.2215/CJN.06430617. Epub 2018 Jan 3. PMID: 29298761; PMCID: PMC5967427.

Mandrekar JN. Receiver operating characteristic curve in diagnostic test assessment. J Thorac Oncol. 2010 Sep;5(9):1315–6. doi: 10.1097/JTO.0b013e3181ec173d. PMID: 20736804.

Maslach C, Jackson SE. The measurement of experienced burnout. J Occupational Behaviour. 1981;2:99–113

Matthews KA, Gump BB, Harris KF, Haney TL, Barefoot JC. Hostile behaviors predict cardiovascular mortality among men enrolled in the multiple risk factor intervention trial. Circulation. 2004;109:66–70

Melamed S, Kushnir T, Shirom A. Burnout and risk factors for cardiovascular diseases. Behav Med. 1992 Summer;18(2):53–60. doi: 10.1080/08964289.1992.9935172. PMID: 1392214.

Melamed S, Shirom A, Toker S, Shapira I. Burnout and risk of type 2 diabetes: a prospective study of apparently healthy employed persons. Psychosom Med. 2006;68:863–869

Pedregosa, F., Varoquaux, G., Gramfort, A., Michel, V., Thirion, B., Grisel, O., Blondel, M., Prettenhofer, P., Weiss, R., Dubourg, V., Vanderplas, J.: Scikit-learn: machine learning in Python. J. Mach. Learn. Res. 12, 2825–2830 (2011)

Robinson MN, Thomas Tobin CS. Is John Henryism a Health Risk or Resource?: Exploring the Role of Culturally Relevant Coping for Physical and Mental Health among Black Americans. J Health Soc Behav. 2021 Jun;62(2):136–151. doi: 10.1177/00221465211009142. PMID: 34100655; PMCID: PMC8370445.

Rolle T, Vue Z, Murray SA, Shareef SA, Shuler HD, Beasley HK, Marshall AG, Hinton A. Toxic stress and burnout: John Henryism and social dominance in the laboratory and STEM workforce. Pathog Dis. 2021 Sep 11;79(7):ftab041. doi: 10.1093/femspd/ftab041. PMID: 34410372; PMCID: PMC8435059.

Rose RJ. Genetic and environmental variance in content dimensions of the MMPI. J Pers Soc Psychol. 1988 Aug;55(2):302–11. doi: 10.1037//0022-3514.55.2.302. PMID: 3171910.

Shirom A, Westman M, Shamai O, Carel R. Effects of work overload and burnout on cholesterol and triglycerides levels: the moderating effects of emotional reactivity among male and female employees. J Occup Health Psychol. 1997;2:275–288.

Smith TW, McGonigle M, Turner CW, Ford MH, Slattery ML. Cynical hostility in adult male twins. Psychosom Med. 1991 Nov-Dec;53(6):684–92. doi: 10.1097/00006842-199111000-00008. PMID: 1758951.

Stewart NH, Arora VM. The Impact of Sleep and Circadian Disorders on Physician Burnout. Chest. 2019 Nov;156(5):1022–1030. doi: 10.1016/j.chest.2019.07.008. Epub 2019 Jul 25. PMID: 31352036; PMCID: PMC6859241.

Teumer A. Common Methods for Performing Mendelian Randomization. Front Cardiovasc Med. 2018 May 28;5:51. doi: 10.3389/fcvm.2018.00051. PMID: 29892602; PMCID: PMC5985452.

Vargas EA, Li Y, Mahalingham R, Hui P, Liu G, Lapedis M, Liu JR. The double edge sword of John Henryism: Impact on patients’ health in the People’s Republic of China. J Health Psychol. 2020 Nov-Dec;25(13-14):2374–2387. doi: 10.1177/1359105318800141. Epub 2018 Sep 19. PMID: 30229675.

Wang X, Trivedi R, Treiber F, Snieder H. Genetic and environmental influences on anger expression, John Henryism, and stressful life events: the Georgia Cardiovascular Twin Study. Psychosom Med. 2005 Jan-Feb;67(1):16–23. doi: 10.1097/01.psy.0000146331.10104.d4. PMID: 15673619.

Whitfield KE, Brandon DT, Robinson E, Bennett G, Merritt M, Edwards C. Sources of variability in John Henryism. J Natl Med Assoc. 2006 Apr;98(4):641–7. PMID: 16623079; PMCID: PMC2569236.

Wong JM, Na B, Regan MC, Whooley MA. Hostility, health behaviors, and risk of recurrent events in patients with stable coronary heart disease: findings from the Heart and Soul Study. J Am Heart Assoc. 2013 Sep 30;2(5):e000052. doi: 10.1161/JAHA.113.000052. PMID: 24080907; PMCID: PMC3835215.

